# In Silico Trial Simulation with Artificial Intelligence-Generated Synthetic Control Cohorts Reproduces Results of a Randomized Controlled Trial in Acute Myeloid Leukemia

**DOI:** 10.64898/2026.07.15.26358123

**Authors:** Kumar Reddy Kakularam, Waldemar Hahn, Susann Winter, Christoph Röllig, Carsten Müller-Tidow, Hubert Serve, Claudia D. Baldus, Lars Fransecky, Christoph Schliemann, Andreas Burchert, Kerstin Schäfer-Eckart, Martin Kaufmann, Johannes Schetelig, Martin Bornhäuser, Jan Moritz Middeke, Jan-Niklas Eckardt

## Abstract

Rising costs, slow accrual and molecular sub-stratification of cancers necessitate novel clinical trial designs. We demonstrate that artificial intelligence-generated synthetic patients can replace real controls to reproduce results of the SORAML trial. Using external multimodal data from 1,377 acute myeloid leukemia (AML) patients from previous trials and a real-world registry, we fine-tuned a tabular foundation model to generate synthetic patients, reproducing clinical and genetic features and outcome associations. Synthetic patients were then matched to the original SORAML intervention group using Cox risk scores, replacing the original control and reproducing the original trial result with near-identical median event-free survival (EFS) and treatment effect (original hazard ratio [HR] 0.64, 95%-confidence interval [CI] 0.47–0.87, *p*=0.004; with synthetic control HR 0.66, 95%-CI 0.48– 0.90, *p*=0.009). Our findings demonstrate that AI-generated synthetic patients can serve as statistically rigorous controls supporting novel trial designs.

## Introduction

Clinical trials in cancer research are too expensive, take too long, and fail too often. The costs for the development of a single cancer drug from bench to market are estimated to exceed one billion US-Dollars with a steady upward trend over the last decade.^1–3^ In addition, drug development in oncology shows devastatingly low success rates, especially for early phase trials.^4–6^ Despite accelerated approval and breakthrough designations shortening time-to-approval for multiple recently approved cancer drugs, development times still span close to a decade in most instances.^7–10^ Slow enrollment frequently delays trial completion, thereby increasing costs and failure rates.^11,12^ Using data from 27 cancer clinical trials, Stewart et al.^13^ estimated that, for every year that is cut from time-to-approval, a median of 79,920 patient-life-years could potentially be saved. These challenges are likely to intensify as precision oncology increasingly stratifies malignancies into smaller, molecularly defined subgroups, thereby shrinking the pool of potentially eligible patients for individual trials. This may lead to multiple trials within the same entity competing for patient enrollment, as patients recruited to a control cohort of one trial could otherwise potentially have been recruited to an investigational cohort in another trial. This may further slow overall cancer drug development times and increase costs as well as failure rates. Hence, novel trial designs are an urgent unmet need. The key question is: What is the alternative to recruiting a standard of care control group?

Generative artificial intelligence (AI) can now produce realistic synthetic datasets across multiple domains, including structured tabular data.^14^ By learning abstract representations of statistical distributions, generative models can produce realistic synthetic patient records that are not mere copies of the underlying originals. In oncology, synthetic tabular data generated from previous clinical trials and real-world registries may enable synthetic control cohorts to augment or potentially replace real controls in prospective clinical trials.^15^ In contrast to historic or real external controls, which are often limited by heterogeneity, missing data, and outdated treatment regimens^16,17^, these synthetic cohorts preserve patient privacy, can be generated at scale, and tailored to fit specific eligibility criteria. We have previously demonstrated that synthetic patient data created by generative AI closely resemble real patients with acute myeloid leukemia (AML). In these synthetic cohorts, disease-related biological correlations and their association with clinical outcomes are preserved, while risks of patient re-identification are effectively mitigated.^18^

In this study, we trained a generative model on AML patient data from previous clinical trials of the German Study Alliance Leukemia (SAL) to generate a synthetic control cohort. We then replaced the original control cohort of the previously completed SORAML trial, a multicenter, randomized, controlled phase 2 trial evaluating the addition of sorafenib to intensive induction therapy.^19^ Our objective was to determine whether a synthetic cohort produced by a generative AI model trained on external multi-source data could reproduce the results and conclusions of a randomized controlled trial (RCT), thereby providing a template for future in silico trial designs in cancer research.

## Methods

### Clinical Trial Data

The phase 2 multicenter RCT SORAML (NCT00893373)^19^ was used to evaluate the feasibility of synthetic control cohort substitution. In SORAML^19^, 267 adult AML patients were randomly assigned to receive two cycles of intensive induction therapy, followed by three cycles of consolidation therapy either with the addition of the multikinase inhibitor sorafenib (n=134) or placebo (n=133), followed by sorafenib or placebo maintenance therapy for 12 months. The primary endpoint was event-free survival (EFS). Events were defined as either primary treatment failure, relapse or death. The final analysis reported improved event-free survival (EFS) with the addition of sorafenib, yet no significant differences in overall survival (OS) were observed.^19^ Importantly, patients from the SORAML trial were not used for synthetic data generation. Synthetic data were generated using data from 1,377 adult AML patients enrolled into three previously conducted RCTs of the SAL (AML96 [NCT00180115]^20^, AML2003 [NCT00180102]^21^, AML60+ [NCT00180167]^22^) and patients from the real-world SAL registry (NCT03188874). Of note, all patients in these trials were treated with intensive induction therapy without the addition of targeted agents (Table S1). Patients with acute promyelocytic leukemia (APL) were not included in any of the previously mentioned trials. All patients gave their written informed consent according to the revised Declaration of Helsinki.^23^ All studies including subsequent retrospective analyses of clinical trial data were previously approved by the Institutional Review Board of the TUD Dresden University of Technology (EK 98032010). Cytogenetic analyses were conducted using standard techniques for chromosome banding and fluorescence-in-situ-hybridization (FISH). Molecular alterations were assessed using Next-Generation Sequencing (NGS) with the TruSight Myeloid Sequencing Panel (Illumina, San Diego, CA, USA). Data preprocessing included removing redundant (age, date of birth) or sparse features, i.e. genetic alterations with frequencies below 2%. Missing values were imputed by using MissForest^24^, a random forest-based imputer, implemented in the HyperImpute Python package^25^. The pre-processed dataset included 51 variables (Table S2).

### Synthetic Tabular Data Generation with TabPFN

Synthetic data generation hinges on a model’s ability to implicitly learn the joint distribution of features observable in the training data and produce novel plausible data based on the same feature distributions^26^. In this study, synthetic tabular patient data was generated using TabPFN (Tabular Prior-Data Fitted Network), a transformer-based model initially introduced by Hollmann et al.^27^ and more recently updated to TabPFN-2.5^28^. Briefly, TabPFN-2.5 is specifically designed for tabular data and trained via meta-learning on a large distribution of tabular datasets, enabling it to approximate Bayesian inference over tabular data distributions in a single forward pass. The synthetic data generation process relies on TabPFN’s ability to implicitly model the joint distribution of features by conditioning on observed data. After fitting the unsupervised wrapper (which internally uses both classification and regression variants of TabPFN), the model samples new data points that are statistically consistent with the training dataset. The preprocessed patient data from AML96, AML2003, AML60+ and real-world SAL registry were divided into a training set of 1,032 patients (∼75%) used for model development and a test set of 345 patients (∼25%) reserved for downstream analysis. The train-test split was applied to isolate model development from performance assessment. This enables an unbiased assessment of the model’s generalization to unseen patient data, rather than merely memorizing patterns from the training data. A total of 10,000 synthetic patients were generated with TabPFN-2.5 using its default configuration and were used for further exploration and validation analysis.

### Risk score-based patient selection

Synthetic data generation algorithms such as TabPFN-2.5 are capable of generating an arbitrary number of synthetic patients. However, to be useful as a control cohort, synthetic patients have to resemble both genotypes and phenotypes of real patients enrolled into the trial to minimize potential confounding by features other than the investigated therapeutic agent. To this end, the available clinical and genetic features were evaluated regarding their impact on EFS using multivariable Cox proportional hazard (CoxPH) models. The patient-specific risk score was calculated as a weighted linear combination of feature values and their corresponding Cox regression coefficients, where larger risk scores corresponded to higher predicted risk and poorer survival outcomes. Only the Cox coefficients of variables that were significantly associated with EFS (*p*<0.05) were included in the risk score calculation (Table S3). Each real patient was subsequently matched to the closest synthetic patient based on risk score similarity, resulting in a one-to-one matching between real and synthetic patient records. Based on this risk stratification, the closest synthetic matches were identified for the test cohort (1:1 matching, n = 345 patients), the SORAML control cohort (1:1 matching, n = 133 patients and 1:2 matching, n = 266 patients), and the SORAML intervention cohort (1:1 matching, n = 134 patients and 1:2 matching, n = 268 patients).

### Statistical analysis

Pairwise statistical analyses were conducted to compare the synthetic control cohort to the real heldout test cohort as well as the real heldout test cohort to the real training cohort. The Shapiro-Wilk test was used to assess normality of continuous variables, indicating non-normal distributions for all continuous variables under investigation. Hence, continuous variables were compared using the Wilcoxon rank sum (syn. Mann-Whitney) test. Categorical variables were compared using Fisher’s exact test. All tests were carried out two-sided. Survival analysis was performed using the Kaplan-Meier method complemented by log-rank tests for differences in time-to-event variables. Univariable Cox regression was used to obtain hazard ratios (HR) and 95% confidence intervals (95%-CI). Statistical significance was determined using a significance level α of 0.05. The Python libraries lifelines^29^ and SciPy^30^ were used for survival and statistical analyses, respectively.

### Performance evaluation

The quality of the generated synthetic data was evaluated using a combination of statistical fidelity, distributional similarity, and survival analysis metrics implemented through the SynthCity framework^31^ and the TabSynDex^32^ evaluation protocol. TabSynDex-based metrics quantify preservation of feature relationships (log-transformed correlation score), distinguishability between real and synthetic samples (propensity mean squared error), coverage of the original data distribution (regularized support coverage), and agreement in summary statistics (basic statistical measure). These metrics are normalized to range between 0 and 1, with values closer to 1 indicating better fidelity between real and synthetic datasets. Distributional similarity was further assessed using Jensen– Shannon distance^33^, Wasserstein distance^34^, and the Kolmogorov–Smirnov statistic^35^, which quantify divergence between real and synthetic data distributions, where lower values indicate better agreement. Furthermore, survival fidelity was evaluated using metrics introduced by Norcliffe et al.^36^ (where values closer to 0 indicate better performance) and implemented in SynthCity^31^: KM divergence score was used to quantify the overall difference between survival curves, optimism score was used to assess if synthetic data overestimates or underestimates the survival outcomes, and short-sightedness score was used to quantify temporal discrepancies, reflecting how well the synthetic data preserves time-dependent survival dynamics.

### Privacy assessment

The generated synthetic patient data was evaluated for privacy preservation and potential memorization. Prior to privacy assessment, synthetic records containing continuous feature values outside the range of the training data were excluded to ensure consistency during distance-based evaluation. For exact match analysis, continuous variables were discretized into 5 quantile-based bins derived from the real training data distribution and synthetic continuous features were subsequently mapped to the corresponding bin intervals. Exact matches between training data and synthetic data were then identified using the combined representation of binary and discretized continuous variables. Further, distance-based privacy metrics, including distance to closest record (DCR)^37^ and nearest neighbor distance (NNDR)^38^ were employed to evaluate similarity and potential memorization. Euclidean distance computed on normalized feature representations was used as a distance measure. For DCR analysis, the distance between each synthetic sample and all real data samples were calculated, and the median of the nearest real sample distance across all synthetic records was reported. Lower DCR values indicate high similarity between synthetic and real samples and may reflect increased memorization. For NNDR analysis, the ratio between the distance to the nearest training sample and the distance to the second-nearest training sample was computed for each synthetic record. The median of these ratios across all records was reported as NNDR. The NNDR value ranges between 0 and 1 and NNDR closer to 0 indicate that a synthetic sample is disproportionately close to a specific real sample, which may suggest potential memorization or privacy leakage. Additionally, membership inference attacks (MIA)^39^ were performed to evaluate whether synthetic samples leaked identifiable information regarding membership in the training dataset. For each real sample, the minimum distance to the synthetic dataset was computed and used as the attack score. Training samples were considered members, whereas test samples were considered non-members. Attack performance was quantified using Receiver Operating Characteristic Area Under the Curve (ROC-AUC) and attack accuracy. ROC-AUC values approaching 0.5 were interpreted as indicating minimal distinguishability between member and non-member samples and therefore limited privacy leakage from the synthetic data generation process. Detailed and elaborate descriptions of the synthetic data privacy evaluation metrics and their implementation and interpretation can be found in the review by Steier et al.^40^

### Code availability

The underlying code generated for the purpose of this study is publicly available at https://github.com/Ai-in-Cancer-org/synnext_aml

## Results

### Synthetic patients mimic genotypes and phenotypes of real patients

TabPFN-2.5 was used to generate synthetic AML patients based on patient data from the AML96^20^, AML2003^21^, AML60+^22^ trials and real-world SAL registry. The combined dataset was divided into a training cohort of 1,032 patients used to fine-tune TabPFN-2.5, a generative foundation model, capturing the underlying data distribution, and a heldout test set of 345 patients for validation. Figure 1 illustrates the workflow of the study. To evaluate faithful mimicry of disease biology and clinical presentation, we compared the distribution characteristics of 31 molecular genetic and ten cytogenetic variables, along with age, sex, and six laboratory values between the test set real patients (that were not used for training the generative model) and both the synthetic patient cohort as well as the real training cohort. Synthetic patients accurately reproduced the feature distributions observed in the real cohort. We did not find any statistically significant differences in age, sex, or laboratory values between real and synthetic patients, except for a lower median platelet count in synthetic patients (46 GPt/l vs. 56 GPt/l, *p*=0.017, Table 1). With regard to genetic alterations, we found only marginal differences in distributions between real test and synthetic patients (Figure 2), wherein only alterations of *KIT* (test 7.2% vs. synthetic 1.7%, *p*=0.001), *RAD21* (test 5.2% vs. synthetic 1.4%, *p*=0.009), *CEBPA* (test 20.0% vs. synthetic 11.0%, *p*=0.002) including biallelic mutations of *CEBPA* (7.0% vs. synthetic 2.9%, *p*=0.021), and extramedullary manifestations of AML (test 16.2% vs. synthetic 10.4%, *p*=0.033) reached statistical significance. Notably, the frequencies of *KIT* (test 7.2% vs. training 4.3%, *p*=0.033) and *RAD21* (test 5.2% vs. training 2.6%, *p*=0.023) alterations also differed between real training and test cohort, reflecting expectable variations between multiple study cohorts.

**Figure 1.**
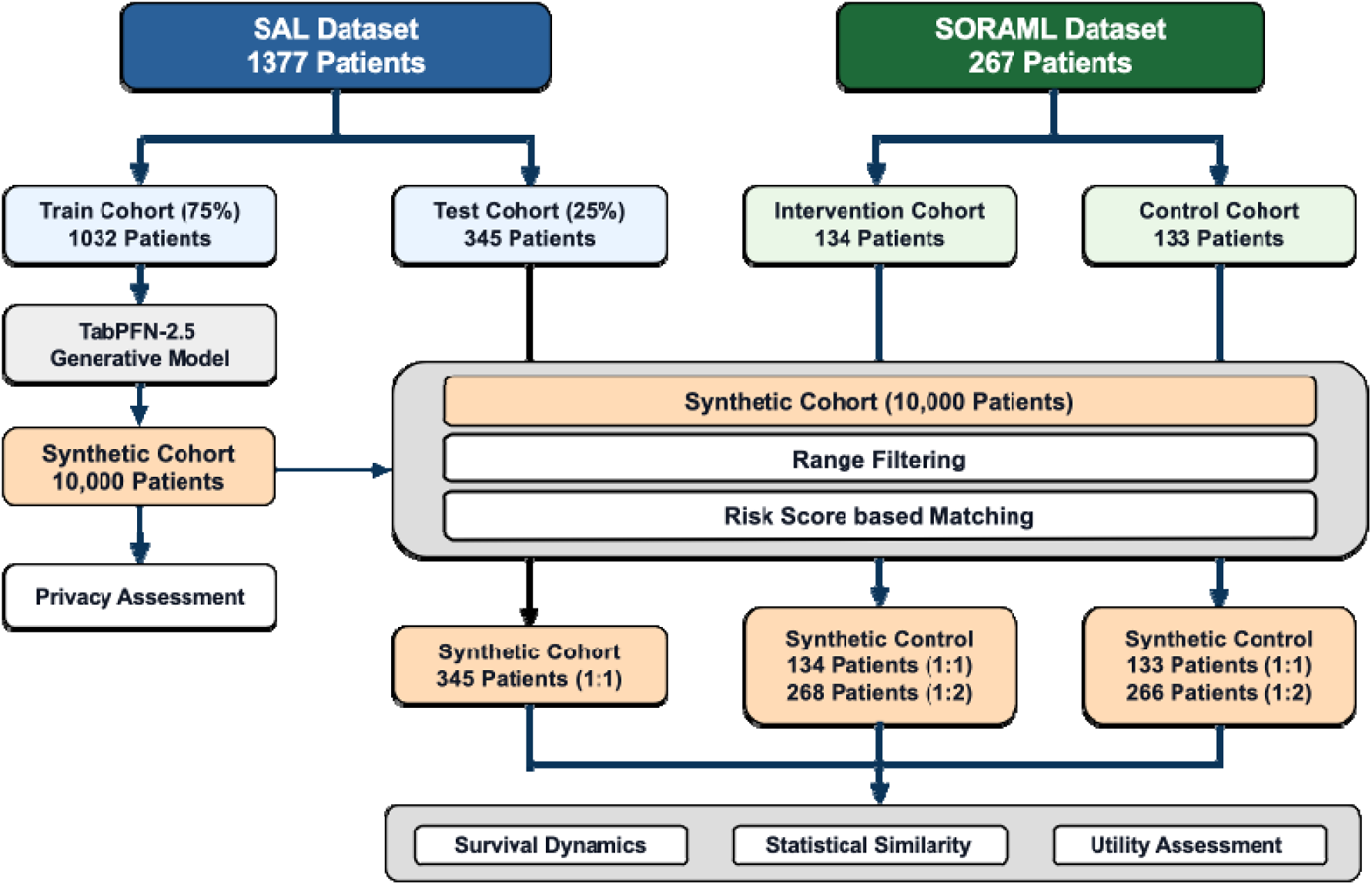
Synthetic data generation and validation workflow. The SAL dataset (n = 1,377) was split into training and test cohorts. The training cohort was used to fine-tune the TabPFN-2.5 generative model and generate a synthetic cohort of 10,000 patients. Synthetic patients were selected using range filtering and risk score-based matching to generate cohorts matched to the SAL test set, the SORAML intervention, or SORAML control arm (1:1 and 1:2 matching), respectively. Statistical similarity, utility, survival dynamics, and privacy assessments were performed to evaluate the quality and fidelity of the generated synthetic data.

**Figure 2.**
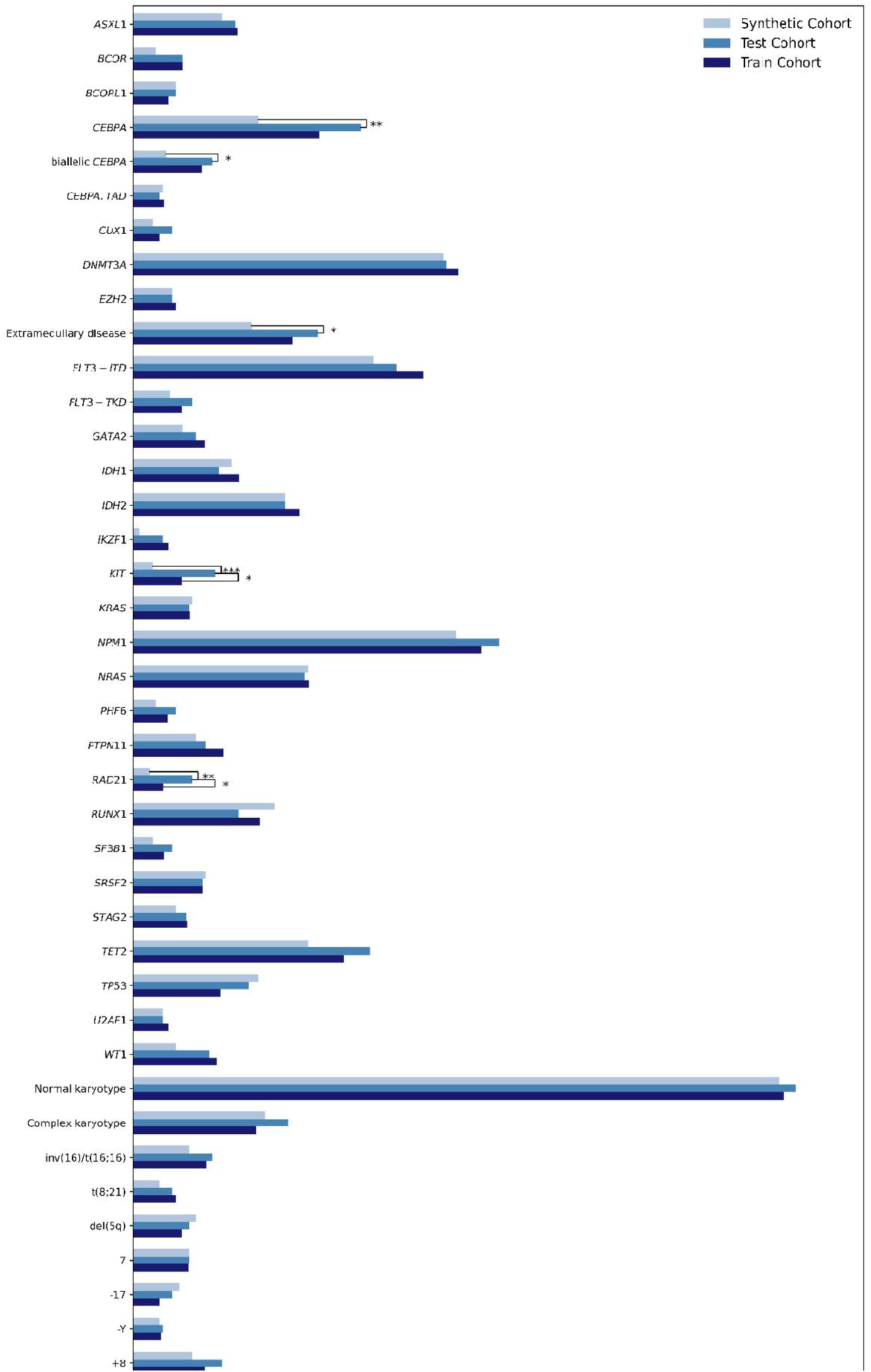
Comparison of genetic alterations between synthetic, real test set, and real training set patients. The distribution of genetic alterations across the 345 real patient test set, 345 matched synthetic patients and 1,032 real training cohort patients is displayed. The percentage of positive cases for 41 genetic and cytogenetic features is shown for each cohort. Features demonstrating statistically significant differences are annotated as follows: *p < 0.05, **p < 0.01, and ***p < 0.001.

**Table 1.** Baseline characteristics. Age, sex and laboratory values are compared between test set real patients with synthetic patients and test set real patients with training set patients. The corresponding *p*-values were computed using Wilcoxon rank sum tests for continuous variables and Fisher’s exact test for sex (categorical).

|  | Real Test | Synthetic Test | *p*-value (Real test Vs Synthetic Test) | Real Train | *p*-value (Real test vs Real Train) |
| --- | --- | --- | --- | --- | --- |
| Number of patients | 345 | 345 |  | 1032 |  |
| Age in years, median (IQR) | 57.0 (46.0 - 66.0) | 57.0 (45.0 - 66.0) | 0.5381 | 58.0 (44.0 - 67.0) | 0.995 |
| White blood cell count in GPt/l, median (IQR) | 21.1 (4.4 - 54.37) | 20.77 (5.15 - 50.84) | 0.967 | 21.25 (5.4 - 58.15) | 0.374 |
| Hemoglobin concentration in mmol/l, median (IQR) | 5.65 (4.84 - 6.5) | 5.71 (4.93 - 6.47) | 0.954 | 5.71 (4.9 - 6.46) | 0.827 |
| Platelet count in GPT/l, median (IQR) | 56.0 (29.0 - 95.0) | 45.71 (24.46 - 80.3) | **0.017** | 48.0 (27.0 - 92.0) | 0.121 |
| LDH in U/l, median (IQR) | 463.0 (282.0 - 886.8) | 442.01 (296.36 - 711.66) | 0.233 | 472.5 (312.0 - 827.25) | 0.855 |
| Bone marrow blast count, median percentage (IQR) | 61.0 (46.5 - 74.0) | 60.53 (44.4 - 76.65) | 0.896 | 62.75 (47.0 - 77.5) | 0.239 |
| Peripheral blood blast count, median % (IQR) | 42.0 (15.0 - 71.0) | 37.24 (14.75 - 70.21) | 0.832 | 41.0 (14.0 - 74.0) | 0.869 |
| Sex n (%) |  |  | 0.402 |  | 0.6188 |
| Female | 159 (46.09) | 171 (49.57) |  | 492(47.67) |  |
| Male | 186 (53.91) | 174 (50.43) |  | 540 (52.33) |  |

### Synthetic data preserve cohort distributions and inter-variable relationships

Comparisons at the individual feature level demonstrated high similarity between real test and synthetic cohorts. To assess similarity at the cohort level, we evaluated a set of complementary metrics for synthetic data evaluation from the TabSynDex^32^ framework capturing distributional alignment, feature space coverage, geometric structure, and inter-feature relationships across all features jointly (Table 2). Basic statistical properties such as mean, median and standard deviation were compared between datasets measured by Basic Statistical Measure (closer to 1 indicates higher similarity), with a score of 0.878 indicating close resemblance. Further, synthetic data closely matched feature probability distributions and empirical cumulative distribution functions of real data, as reflected by a low Jensen–Shannon distance (0.005; closer to 0 indicates greater similarity) and a high Kolmogorov– Smirnov statistic (0.973; closer to 1 indicates greater similarity). Coverage of the real data space was measured using Regularized Support Coverage Score, yielding a value of 0.903 (closer to 1 indicates alignment of plausible feature ranges), reflecting that most regions of the empirical feature space were represented in the synthetic cohort. Consistent with this, the Wasserstein distance (1.295; lower values indicate closer alignment) suggested a similar shape in the overall geometry of the data. Beyond marginal distributions, synthetic data preserved relationships between variables, with a Log-Transformed Correlation Score of 0.701 (1 indicates perfect correlation). A logistic regression classifier trained to distinguish between real and synthetic samples yielded a Propensity Mean Squared Error of 0.778 (closer to 1 indicates an arbitrary classification), demonstrating that a trained classifier could not reliably distinguish real and synthetic samples. Importantly, across all evaluated metrics, similarity between synthetic and real test data was comparable to that observed between real test and training data (Table 2), supporting the fidelity of the synthetic cohort.

**Table 2.** Performance metrics comparisons between 345 patients from the test cohort, 345 synthetic patients, and 1,032 patients from the training cohort.

| Fidelity Metrics | Range (inference) | Real Test vs Synthetic Test | Real Test vs Real Train |
| --- | --- | --- | --- |
| Log-transformed Correlation Score | 0 to 1 (closer to 1 is better) | 0.701 | 0.683 |
| Propensity Mean Squared Error | 0 to 1 (closer to 1 is better) | 0.778 | 0.761 |
| Regularized Support Coverage | 0 to 1 (closer to 1 is better) | 0.903 | 0.954 |
| Basic Statistical Measure | 0 to 1 (closer to 1 is better) | 0.878 | 0.934 |
| Jensen–Shannon Distance | 0 to 1 (closer to 0 indicates identical distributions) | 0.005 | 0.003 |
| Wasserstein Distance | 0 to infinity (lower means closer) | 1.295 | 1.221 |
| Kolmogorov–Smirnov statistic | 0 to 1 (closer to 1 means the distributions are identical) | 0.973 | 0.983 |

### Synthetic patients replicate outcome and survival characteristics of real patients

The preservation of baseline characteristics and inter-variable relationships translated into the preservation of outcome dynamics in synthetic patients. Comparing median EFS between real test set patients and synthetic patients showed no statistically significant differences (real test set: 7.33 months, interquartile range [IQR] 1.12 - 25.81 months vs. synthetic: 6.11 months, IQR 1.38 - 22.72 months, *p*=0.852). In accordance, there were no significant differences in median EFS between real test and real training set patients (real training set: 6.71 months, IQR 1.08 - 28.41 months, *p*=0.748).

Despite non-significant differences in median EFS between real and synthetic patients, outcome kinetics may still differ as generative models may potentially fail to appropriately capture events, censoring or stabilization of the survival curve. In our analysis, we found synthetic patients to faithfully resemble survival kinetics of real patients over a median and maximum follow up period of 156 and 163 months, respectively. Kaplan-Meier survival graphs indicated a close resemblance between real test and synthetic patient outcomes (compared to real test and real training set), with no significant differences between the two (log-rank *p*=0.807, Figure 3). Accordingly, KM divergence score - a measure for proximity or separation between survival curves - showed close agreement between real test and synthetic patients at a value of 0.014 (closer to 0 is better). Further, synthetic data did not exhibit overt optimism or pessimism, i.e. under-or overestimating the occurrence of events throughout the follow-up period, reflected in an optimism score of −0.011 (closer to 0 is better). This was supported by low short-sightedness of 0.043 (closer to 0 is better), indicating the model adequately captured survival behavior even during later stages of the follow-up period when training data was increasingly sparse. Lastly, censoring behavior was similar between synthetic and real test set patients throughout the follow-up period.

**Figure 3.**
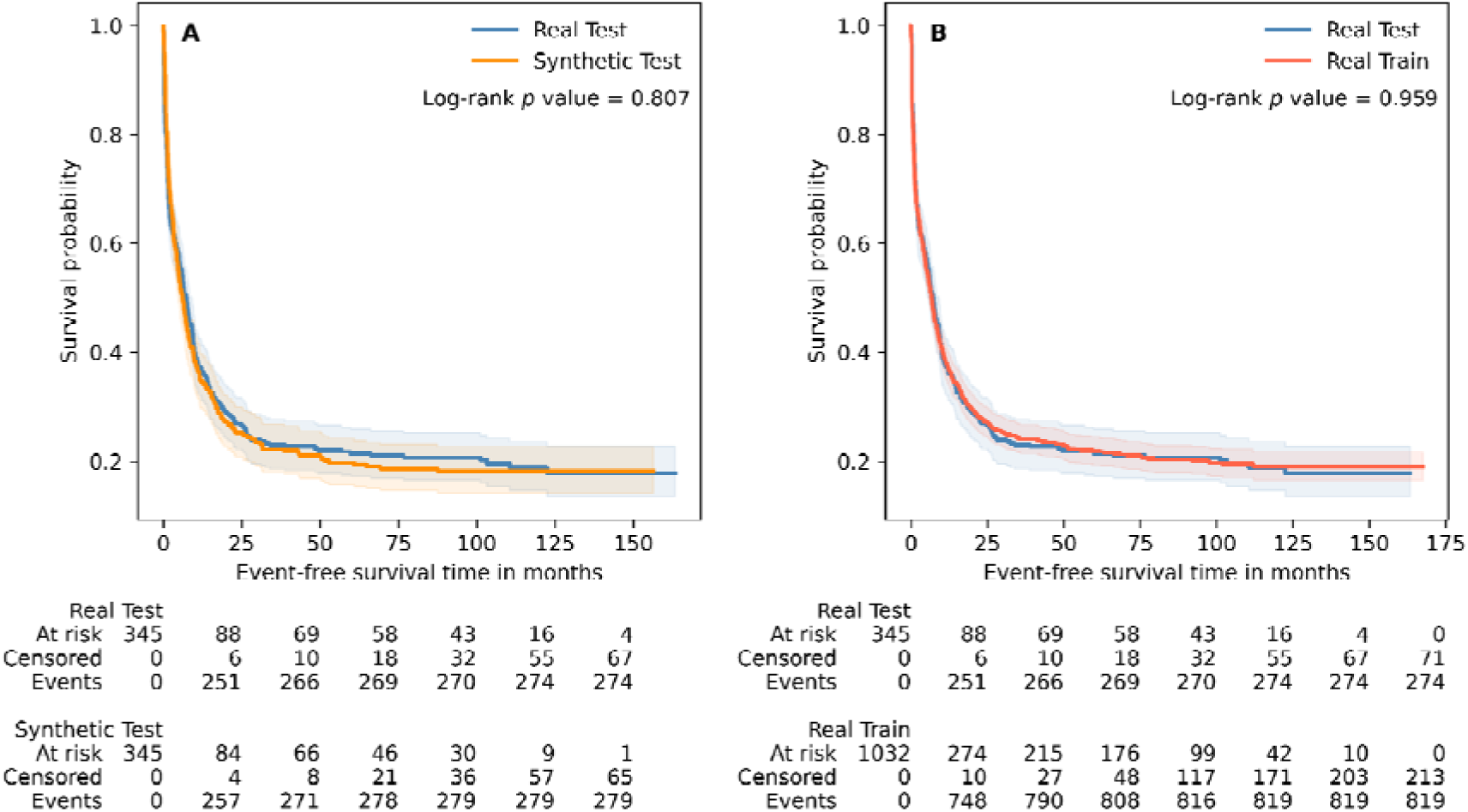
Synthetic patients capture survival dynamics of real patients. Kaplan–Meier survival curves with corresponding risk tables and log-rank *p*-values comparing 345 patients from the test cohort with (A) 345 synthetic patients or (B) 1,032 patients from the training cohort. Risk tables display the number of patients at risk and censored events at each time point for the respective cohorts.

### Synthetic data preserves clinically relevant disease-specific characteristics

AML is a biologically heterogenous disease characterized by diverse genetic alterations affecting patient outcome.^41^ Additionally, increasing age is associated with a higher incidence of AML, is an important determinant of prognosis, modifies the effects of molecular and cytogenetic abnormalities, and influences treatment selection.^42,43^ To be clinically meaningful, synthetic patient data must therefore preserve both the direction (favorable or unfavorable) and magnitude of associations between disease-specific features and patient outcomes. To assess this, we compared outcomes in synthetic and real patients across clinically relevant prognostic subgroups (Figure 4). Favorable-risk features included *NPM1* mutations, core-binding factor (CBF) leukemias, and normal karyotypes (NK), whereas adverse-risk features included *TP53* alterations, complex karyotypes (CK), and older age. Across all subgroups, synthetic patients closely reproduced the survival patterns observed in real patients. No significant differences in EFS were found between synthetic and real patients within any subgroup defined by the presence or absence of any of these features. Vice versa, a given feature still remained associated with EFS, irrespective of whether it was found in real or synthetic patients.

**Figure 4.**
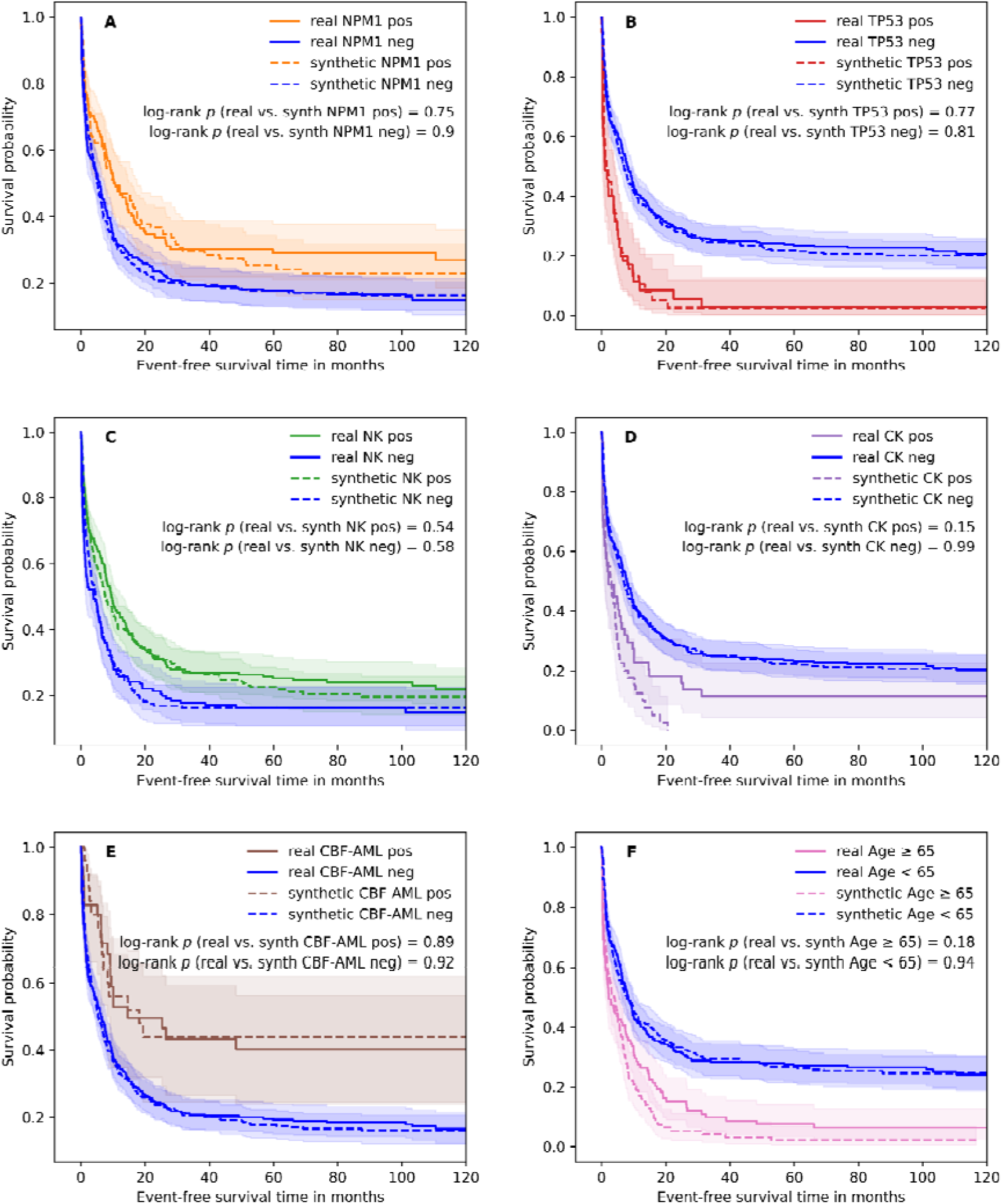
Survival dynamics of real and synthetic patient cohorts in prognostically relevant AML subgroups. Kaplan–Meier survival curves are shown for patients stratified by: (A) *NPM1* mutation status, (B) *TP53* mutation status, (C) normal karyotype (NK), (D) complex karyotype (CK), (E) core-binding factor leukemias (CBF-AML), and (F) age categorized using a 65-year cutoff. Universally, the effect size and significance level of prognostically relevant markers was preserved between real and synthetic patients. Alterations of *NPM1*, normal karyotypes (NK) and core-binding-factor leukemias are established markers of favorable outcomes, whereas alterations of *TP53*, complex karyotypes (CK) and older age are established markers of adverse outcomes. No significant differences in event-free survival were observed between real and synthetic patients within any subgroup defined by the presence or absence of these features, indicating that synthetic patients reproduced subgroup-specific survival patterns. At the same time, each feature retained its expected prognostic association in both real and synthetic cohorts, demonstrating preservation of clinically relevant relationships between disease characteristics and outcome.

### Synthetic patients can be used as controls in clinical trials

Our previous evaluations of synthetic patients trained on AML96, AML2003, AML60+ and SAL real-world registry data demonstrated high fidelity and concordance in survival metrics compared to a heldout test set derived from these same studies. To evaluate downstream usability of synthetic patients, we used an external clinical trial cohort derived from the phase 2 SORAML trial, comprising 133 control and 134 intervention patients that were not part of model training or testing.

A Cox proportional hazards model was used to estimate associations between clinical and molecular covariates and event-free survival (EFS). Regression coefficients from this model were then used to derive individualized risk scores representing weighted combinations of prognostic features (Table S3). Synthetic patients were selected by matching these risk scores between real and synthetic cohorts. Compared with random sampling, this risk-score-based approach selected synthetic patient control cohorts with substantially greater clinical and molecular similarity to real patients and reproduced corresponding survival dynamics far more reliably (99% versus 32% of cases across 133 runs).

We evaluated two scenarios: Using risk score-based matching, (i) synthetic patients were matched to the original SORAML control group and (ii) synthetic patients were matched to the original SORAML intervention group. The first scenario evaluated comparability to the original control cohort, while the second scenario simulates real-world addition of synthetic controls to a prospective RCT where only a real intervention arm exists. As a baseline, we compared the original intervention and control groups of the SORAML trial, where median EFS differed significantly between real intervention and real control, yielding significantly improved EFS with the addition of sorafenib (25.38 vs. 9.80 months, HR 0.64, 95%-CI 0.47-0.87, *p*=0.004).

In the first scenario, 133 synthetic patients were matched to the original 133 control group patients and subsequently replaced the original control cohort. Median EFS was almost identical between the original SORAML control cohort compared to the synthetic control cohort (9.80 vs 9.86 months, HR 0.87, 95%-CI 0.65-1.16, *p*=0.335). Comparing the original SORAML intervention cohort to this synthetic control cohort, the favorable effect of sorafenib was preserved (25.38 vs. 9.86 months, HR 0.74, 95%-CI 0.54-1.02, *p*=0.066), yet statistical significance was narrowly missed (Figure 5A). In clinical research, this result may lead to the assumption that the study may have been underpowered. To test this, we doubled the size of the synthetic control cohort (n=266), yielding again a median EFS of 9.86 months for the synthetic patients, however, now also showing a statistically significant difference between original intervention cohort and the double-sized synthetic control in this 1:2 comparison (HR 0.73, 95%-CI 0.55-0.96, *p*=0.024; Figure 5B).

**Figure 5.**
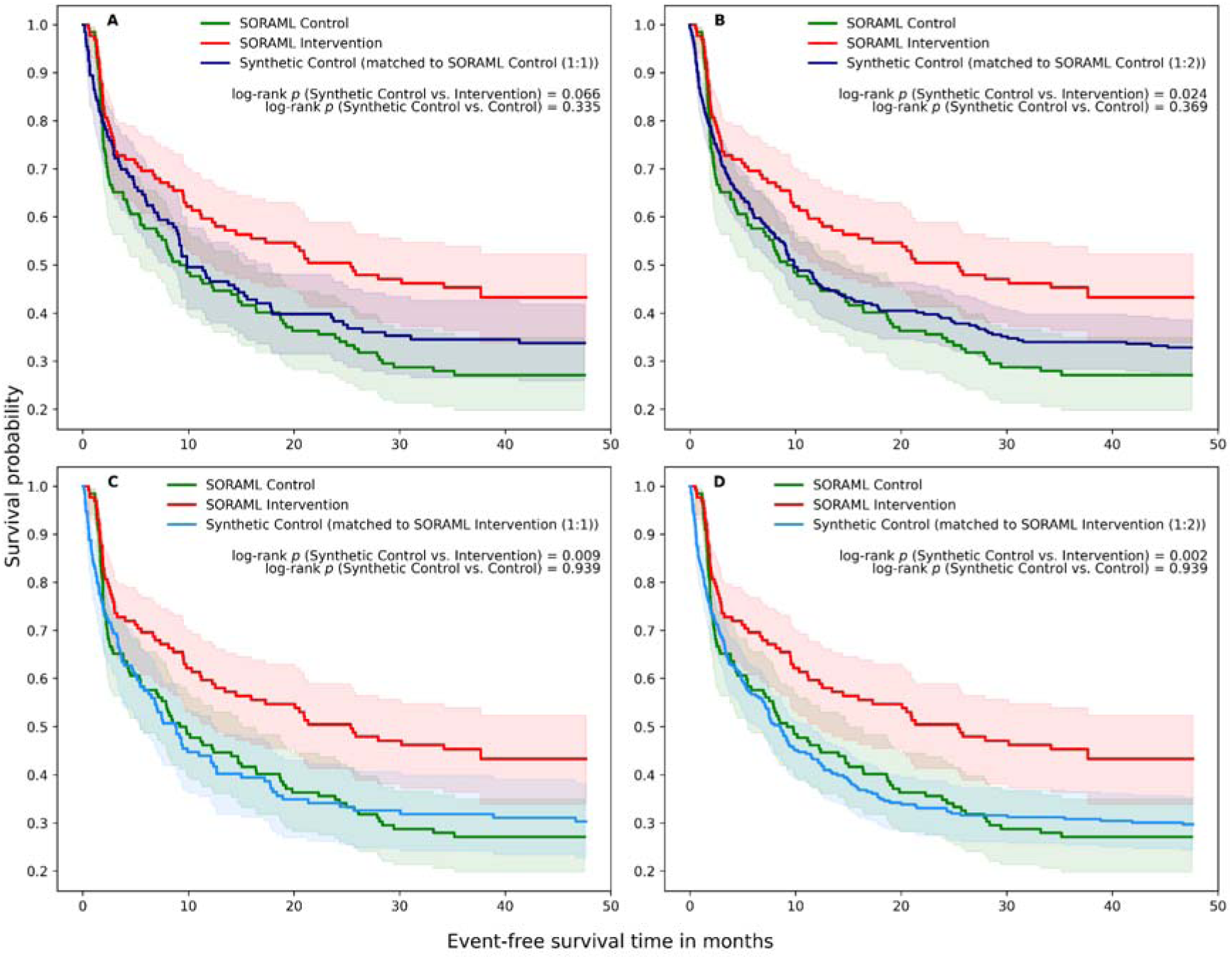
Synthetic controls reproduce findings of the original SORAML trial. Survival dynamics of the original SORAML control (133 patients) and SORAML intervention group (134 patients) are compared with (A) a synthetic control cohort matched to the original SORAML control cohort in a 1:1 ratio (133 patients each) or (B) a 1:2 ratio (266 synthetic control patients), and (C) a synthetic control cohort matched to the original SORAML intervention in a 1:1 ratio (134 patients each) or (D) a 1:2 ratio (268 synthetic control patients). When matching synthetic patients to the original SORAML intervention group, both effect size and significance level of the original trial were reproduced irrespective of control group size (C, D). When matching synthetic patients to the original SORAML control group, the 1:1 setup preserved original effect directionality, but slightly missed conventional statistical significance (A). However, increasing the synthetic control group size in this scenario in a 1:2 ratio, effect size was preserved while also reaching statistical significance (B).

In the second scenario imitating the most probable real-world implementation of a synthetic control arm, 134 synthetic patients were matched to the original 134 intervention group patients and subsequently replaced the original control cohort. Again, median EFS did not differ significantly between the original SORAML control cohort compared to this synthetic control cohort (9.80 vs. 8.78 months, HR 0.99, 95%-CI 0.74-1.32, *p*=0.939), while median EFS differed significantly between original SORAML intervention compared to this synthetic control cohort (25.38 vs. 8.78 months, HR 0.66, 95%-CI 0.48-0.90, *p*=0.009, Figure 5C). Doubling the size of the synthetic control group again showed a median EFS of 8.47 months and essentially reproduced the effect size and significance level observed in the 1:1 comparison (HR 0.65, 95%-CI 0.49-0.85, *p*=0.002; Figure 5D).

### Synthetic data preserves patient privacy

To ascertain training set patient privacy, the generated set of 10,000 synthetic patients was evaluated using exact match analysis, distance to closest record^37^, nearest neighbour distance ratio^38^, and membership inference attacks^39^. Exact match analysis did not identify training-set-to-synthetic matches. Distance-based analyses also indicated no memorization or privacy leakage, concordantly observed with a median of 1.94 for Euclidean distance to closest record (where values close to zero raise privacy concerns) and a median nearest neighbor distance ratio of 0.95 (where values closer to one indicate better privacy preservation). For membership inference attacks, the closest synthetic record was identified for both member samples (training data) and non-member samples (holdout test data), evaluating whether inclusion of a sample in the training data could be inferred from synthetic data. Attack performance was random (AUROC 0.557; accuracy 0.524), indicating minimal distinguishability between samples. Together, these analyses showed no evidence that the generative model memorized individual patients or leaked meaningful information about training-set membership.

## Discussion

We demonstrated that AI-generated synthetic patients can be used as synthetic controls to reproduce the results of a randomized controlled trial. We trained a generative model on data of AML patients treated with non-targeted standard of care chemotherapy. The resulting synthetic patients reproduced disease biology and clinical presentation of real AML patients, preserved associations between both genetic and clinical features with outcome and replicated long-term survival patterns. We then matched synthetic patients to fit the phase 2 randomized controlled trial SORAML, replacing the original control cohort and simulating a prospective one-armed trial design with an added synthetic control group. Matching synthetic patients to the intervention cohort in a 1:1 ratio reproduced the treatment effect observed in the original SORAML trial^19^, yielding a nearly identical hazard ratio and a statistically significant improvement in event-free survival consistent with the original trial report. When synthetic patients were instead matched to the original control cohort, the estimated treatment effect remained virtually unchanged in both direction and magnitude, but narrowly missed conventional statistical significance. In such a scenario in clinical research, concerns about limited statistical power are commonly voiced. Doubling the size of this synthetic control group (matched to original control patients) delivered virtually the same median EFS and hazard ratio, however now resulting in a statistically significant finding. Our results suggest that synthetic patients may therefore not only replace or augment conventional control groups, but could also increase statistical power for existing trials and help resolve borderline findings in potentially underpowered settings. Ultimately, synthetic control cohorts may provide access to investigational therapies for more patients and allow for faster recruitment across the portfolio of trials, thereby enabling patients to partake in the advancements of precision oncology rather than being randomized to often outdated standard-of-care control arms.

While previous studies on synthetic data frequently assessed fidelity by comparing real and synthetic samples using statistical metrics that indicate feature distribution alignment^15^, statistical mimicry is necessary but not sufficient. Clinical application requires demonstrating usability, for instance, by enabling meaningful clinical inference in a trial setting such as ours where synthetic patients not only mimicked biology and clinical presentation, but accurately reproduced trial findings. Unlike conventional external controls, synthetic cohorts reduce privacy concerns, can be tailored to fit the trial in need and generated at scale. The latter is particularly important in rare diseases where multiple trials may compete for the same limited pool of patients eligible for enrollment. Synthetic controls may therefore enable trials (especially in rare diseases) to fully enroll patients in intervention groups, allowing more patients to receive novel treatment options while potentially accelerating trial accrual and cutting costs.

Several limitations have to be acknowledged. First, synthetic data are no panacea as they are limited by the underlying generative model and training data. Especially training data pose a constraint on synthetic data generation as a sizable training cohort is required, introducing a conundrum of needing a large real training cohort to produce a large synthetic cohort in the first place. Importantly, patient populations, biological subgroups and treatment effects not represented in the training data cannot be synthesized, yet, biases present in the training data may be propagated into the generated data.^44,45^ In our study, training data were derived from intensively treated adult AML patients at German academic centers before the adoption of targeted therapies, which may limit applicability of our synthetic dataset to other populations and trial settings. Limited representativeness and potentially propagated biases have to be considered when using synthetic data in the context of clinical trials that may ultimately lead to the approval of a novel drug, device, or intervention and properties of the underlying training data should ideally be disclosed. Hence, we publicly release our full synthetic dataset of 10,000 patients for downstream analyses by the scientific community. Finally, synthetic data reduce risks of patient re-identification, yet they are not inherently privacy-proof. Before data sharing and downstream use, synthetic patients should be assessed for privacy preservation. In our analysis, we did not find exact copies of training data patients and synthetic patients were sufficiently dissimilar from training data patients, effectively protecting patient privacy. While we aimed at simulating the implementation of a synthetic control cohort in a one-armed trial setting, where investigators only have access to the fully recruited intervention cohort, our study does not constitute a real prospective synthetically-controlled design. Future work will focus on three-armed trials (synthetic control, real control, real intervention) to prospectively validate the usability of synthetic controls.

In summary, we demonstrated that AI-generated synthetic patients mimic disease biology as well as clinical presentation and can reproduce the conclusions of a randomized clinical trial, supporting synthetic data generation as a tool for clinically meaningful inference and a potential backbone of future trial designs.

## Data Availability

The synthetic datasets generated and analyzed for the purpose of this study are publicly available at https://zenodo.org/records/20702693 or via https://doi.org/10.5281/zenodo.20702693

## Acknowledgements

This study was supported in part by Else Kröner-Fresenius-Stiftung (Grant number 2024_EKEA.146) and Deutsche Krebshilfe (German Cancer Aid, Project Number 70117128). The authors gratefully acknowledge the computing time made available to them on the high-performance computer at the NHR Center of TU Dresden. This center is jointly supported by the Federal Ministry of Research, Technology and Space of Germany and the state governments participating in the NHR. We thank all contributing physicians, laboratories, and nurses associated with the German Study Alliance Leukemia and especially the participating patients for their valuable contributions.

## Authorship Contributions

**Kumar Reddy Kakularam:** conceptualization (equal); data curation (equal); formal analysis (lead); investigation (equal); methodology (equal); resources (equal); software (lead); validation (equal); visualization (lead); writing—original draft preparation (equal). **Waldemar Hahn**: conceptualization (supporting); data curation (supporting); formal analysis (equal); investigation (equal); methodology (supporting); resources (supporting); software (supporting); validation (equal); writing—review and editing (equal). **Susann Winter**: data curation (supporting); formal analysis (supporting), investigation (equal), validation (equal), writing—review and editing (equal). **Christoph Röllig**: data curation (supporting); formal analysis (supporting); investigation (supporting); resources (supporting), validation (supporting); writing—review and editing (equal). **Carsten Müller-Tidow:** data curation (supporting); formal analysis (supporting); investigation (supporting); resources (supporting), validation (supporting); writing—review and editing (equal). **Hubert Serve**: data curation (supporting); formal analysis (supporting); investigation (supporting); resources (supporting), validation (supporting); writing—review and editing (equal). **Claudia Baldus**: data curation (supporting); formal analysis (supporting); investigation (supporting); resources (supporting), validation (supporting); writing—review and editing (equal). **Lars Fransecky:** data curation (supporting); formal analysis (supporting); investigation (supporting); resources (supporting), validation (supporting); writing—review and editing (equal). **Christoph Schliemann:** data curation (supporting); formal analysis (supporting); investigation (supporting); resources (supporting), validation (supporting); writing—review and editing (equal). **Andreas Burchert:** data curation (supporting); formal analysis (supporting); investigation (supporting); resources (supporting), validation (supporting); writing—review and editing (equal). **Kerstin Schäfer-Eckart:** data curation (supporting); formal analysis (supporting); investigation (supporting); resources (supporting), validation (supporting); writing—review and editing (equal). **Martin Kaufmann:** data curation (supporting); formal analysis (supporting); investigation (supporting); resources (supporting), validation (supporting); writing—review and editing (equal). **Johannes Schetelig**: data curation (supporting); formal analysis (supporting); investigation (supporting); resources (supporting), validation (supporting); writing—review and editing (equal). **Martin Bornhäuser**: funding acquisition (equal); data curation (supporting); formal analysis (supporting); investigation (equal); resources (supporting), validation (equal); writing—review and editing (equal). **Jan Moritz Middeke**: conceptualization (supporting), funding acquisition (equal); data curation (supporting); formal analysis (supporting); investigation (equal); resources (equal), validation (equal); writing—review and editing (equal). **Jan□Niklas Eckardt**: Conceptualization (lead); funding acquisition (equal); data curation (equal); formal analysis (equal); investigation (equal); methodology (equal); project administration (lead); resources (equal); software (supporting); validation (equal); visualization (supporting); writing—original draft preparation (equal).

