## Supplemental Data for "In Silico Trial Simulation with Artificial Intelligence-Generated Synthetic Control Cohorts Reproduces Results of a Randomized Controlled Trial in Acute Myeloid Leukemia"

### Supplements

**Table S1.** Summary of trial regimens

| trial name | clinicaltrials.gov identifier | trial duration | protocol summary |
|---|---|---|---|
| AML96 | NCT00180115 | 1996-2008 | risk-adapted postremission treatment regarding allogeneic stem cell transplantation for high-risk AML and related allogeneic and autologous stem cell transplantation for standard-risk AML, and randomization between intermediate-dose and high-dose cytarabine within the first post-remission course |
| AML2003 | NCT00180102 | 2003-2009 | early allogeneic stem cell transplantation in post-induction aplasia for high-risk AML, factorial design with four therapy arms with two factors of two stages (intensified vs. standard therapy and cytarabine vs. cytarabine + mitoxantrone + amsacrin) |
| AML60+ | NCT00180167 | 2005-2010 | Patients ≥ 60 years, mitoxantron on day 1,2,3 + cytarabine on days 1,3,5,7 vs. DA 7+3 |

| | | | |
|---|---|---|---|
| SORAML | NCT00893373 | 2011-2014 | Standard therapy + sorafenib vs. standard therapy + placebo |
| SAL bioregistry | NCT03188874 | 2010-present | Prospective registry of AML patients |

**Table S2.** Demographic values, laboratory values, outcome variables, genetic, and cytogenetic markers included in the datasets.

| Patient Variables | Data Type | Units/Values |
|---|---|---|
| **Demographic variables** | | |
| Age | Continuous | In years |
| Sex | Binary | 1=Male; 0=Female |
| **Laboratory values** | | |
| White blood cell count | Continuous | GPT/l |
| Hemoglobin concentration | Continuous | mmol/l |
| Platelet count | Continuous | GPT/l |
| LDH (Lactate Dehydrogenase) concentration | Continuous | U/l |
| Bone marrow blast count | Continuous | Count |
| Peripheral blood blast count | Continuous | Count |
| **Outcome variables** | | |
| Event-Free survival time | Continuous | In Months |
| Event-Free survival status | Binary | 1=Event occurred; 0=Censored |
| **Genetic markers** | | |
| *ASXL1* | Binary | 1=Mutation present; 0=Mutation absent |
| *BCOR* | Binary | 1=Mutation present; 0=Mutation absent |
| *BCORL1* | Binary | 1=Mutation present; 0=Mutation absent |
| *CEBPA* | Binary | 1=Mutation present; 0=Mutation absent |
| *biallelic CEBPA* | Binary | 1=Mutation present; 0=Mutation absent |

| | | |
|---|---|---|
| *CEBPA.TAD* | Binary | 1=Mutation present; 0=Mutation absent |
| *CUX1* | Binary | 1=Mutation present; 0=Mutation absent |
| *DNMT3A* | Binary | 1=Mutation present; 0=Mutation absent |
| Extramedullary disease (EXAML) | Binary | 1=Mutation present; 0=Mutation absent |
| *EZH2* | Binary | 1=Mutation present; 0=Mutation absent |
| *FLT3*-ITD | Binary | 1=*FLT3*-ITD present; 0=*FLT3*-ITD absent |
| *FLT3-TKD* | Binary | 1=Mutation present; 0=Mutation absent |
| *GATA2* | Binary | 1=Mutation present; 0=Mutation absent |
| *IDH1* | Binary | 1=Mutation present; 0=Mutation absent |
| *IDH2* | Binary | 1=Mutation present; 0=Mutation absent |
| *IKZF1* | Binary | 1=Mutation present; 0=Mutation absent |
| *KIT* | Binary | 1=Mutation present; 0=Mutation absent |
| *KRAS* | Binary | 1=Mutation present; 0=Mutation absent |
| *NPM1* | Binary | 1=Mutation present; 0=Mutation absent |
| *NRAS* | Binary | 1=Mutation present; 0=Mutation absent |
| *PHF6* | Binary | 1=Mutation present; 0=Mutation absent |
| *PTPN11* | Binary | 1=Mutation present; 0=Mutation absent |
| *RAD21* | Binary | 1=Mutation present; 0=Mutation absent |
| *RUNX1* | Binary | 1=Mutation present; 0=Mutation absent |
| *SF3B1* | Binary | 1=Mutation present; 0=Mutation absent |
| *SRSF2* | Binary | 1=Mutation present; 0=Mutation absent |
| *STAG2* | Binary | 1=Mutation present; 0=Mutation absent |
| *TET2* | Binary | 1=Mutation present; 0=Mutation absent |
| *TP53* | Binary | 1=Mutation present; 0=Mutation absent |
| *U2AF1* | Binary | 1=Mutation present; 0=Mutation absent |
| *WT1* | Binary | 1=Mutation present; 0=Mutation absent |

| Cytogenetic Markers | | |
|---|---|---|
| Normal karyotype (NK) | Binary | 1 = present; 0 = absent |
| Complex karyotype (CK) | Binary | 1 = present; 0 = absent |
| inv(16)/t(16;16) | Binary | 1 = present; 0 = absent |
| t(8;21) | Binary | 1 = present; 0 = absent |
| del(5q) | Binary | 1 = present; 0 = absent |
| -7 | Binary | 1 = present; 0 = absent |
| -17 | Binary | 1 = present; 0 = absent |
| -Y | Binary | 1 = present; 0 = absent |
| +8 | Binary | 1 = present; 0 = absent |
| +21 | Binary | 1 = present; 0 = absent |

**Table S3.** Hazard ratios, regression coefficients, and associated p-values for covariates, demonstrating a statistically significant association with event-free survival (EFS) in Cox proportional hazard analysis. Coefficients represent the log hazard ratios estimated by the model, where positive values indicate increased risk and negative values indicate reduced risk associated with EFS.

| **Patient Variables** | **Cox co-efficient** | ***Hazard Ratio*** | ***p* value** |
|---|---|---|---|
| Age | 0.6164 | 1.8522 | 0.0000 |
| White blood cell count | 0.1127 | 1.1193 | 0.0002 |
| *ASXL1* | 0.3873 | 1.4730 | 0.0005 |
| *BCOR* | 0.3283 | 1.3886 | 0.0291 |
| *CEBPA* | -0.4093 | 0.6641 | 0.0002 |
| *CEBPA.TAD* | 0.8803 | 2.4116 | 0.0000 |
| *DNMT3A* | 0.2129 | 1.2373 | 0.0053 |
| Extramedullary disease (EXAML) | 0.2621 | 1.2997 | 0.0040 |
| *IDH2* | 0.2264 | 1.2541 | 0.0169 |
| *IKZF1* | 0.4479 | 1.5650 | 0.0158 |

| | | | |
|---|---|---|---|
| *NPM1* | -0.6170 | 0.5396 | 0.0000 |
| *PTPN11* | 0.2627 | 1.3004 | 0.0292 |
| *SF3B1* | 0.4091 | 1.5054 | 0.0274 |
| *SRSF2* | 0.3471 | 1.4150 | 0.0083 |
| *TET2* | 0.1636 | 1.1777 | 0.0425 |
| *TP53* | 0.6042 | 1.8298 | 0.0002 |
| -7 | 0.6055 | 1.8322 | 0.0001 |
| -Y | -0.4787 | 0.6196 | 0.0306 |
